# Generalizable mapping of the Epileptogenic Zone from interictal Intracranial EEG using an implant topology-aware Transformer

**DOI:** 10.64898/2026.11.23.26368567

**Authors:** Petr Nejedlý, Petr Klimeš, Václav Křemen, Radek Janča, Barbora Matoušková, Valentina Hrtoňová, Filip Mívalt, Kamila Lepková, Petr Ježdík, Martin Pail, Pavel Daniel, Irena Doležalová, Derek Southwell, Lara Wadi, Kassem Jaber, Pilar Bosque-Varela, Jeff Hall, Raluca Pana, Pavel Jurák, Petr Marusič, Pavel Kršek, Gregory Worrell, Birgit Frauscher, Milan Brázdil

## Abstract

Presurgical localization of the epileptogenic zone remains a central challenge in drug-resistant focal epilepsy. At the same time, the abundant interictal intracranial EEG (iEEG) data routinely acquired during presurgical evaluation remain an underexploited source of localizing information. We developed an iEEG implant topology-aware transformer that combines interictal iEEG features with electrode coordinates to predict resection targets. The model was trained exclusively on three public datasets (n = 476) and tested on cohorts from five independent centers across three countries (n = 216). In patients with favorable surgical outcomes (Engel I; n = 120), the median area under the precision–recall curve (AUPRC) was 0.71, and the area under the receiver operating characteristic curve (AUROC) was 0.92, outperforming interictal spike rate and clinically defined seizure-onset zone baselines in patients with favorable outcomes (P < 0.05 for both AUPRC and AUROC). Patients with unfavorable outcomes (Engel II–IV; n = 96) had lower prediction–resection alignment (median AUPRC, 0.42), and their highest-confidence predictions were located farther from resected tissue (P = 0.005). These findings establish a generalizable framework that integrates neural activity and brain topology to identify epileptogenic tissue from interictal recordings, potentially improving surgical success rates and reducing reliance on resource-intensive seizure capture.

## Introduction

Epilepsy affects approximately 70 million people worldwide^1^, with approximately one-third of patients suffering from seizures resistant to antiseizure medications^2^. For these patients, resection or ablation of the epileptogenic zone (EZ) offers the best prospect of seizure freedom, with reported success rates of 50–70%^3^. The EZ is commonly defined as the tissue whose treatment is necessary and sufficient to achieve seizure freedom; however, it is a theoretical construct that cannot be directly observed^4^. Its delineation is therefore inferred by integrating electrophysiological, imaging, and clinical evidence. As surgical candidates increasingly present with complex epilepsies, this process has shifted from identifying a discrete epileptic focus towards characterizing distributed epileptogenic networks^5^. Invasive intracranial electroencephalography (iEEG) investigations consequently often employ extensive implantation schemes comprising up to 256 channels in commercially available standard clinical amplifiers. The resulting volume and complexity of the data challenge expert interpretation, particularly during interictal periods, and motivate computational methods capable of detecting subtle abnormalities and modeling interactions across the implanted network^6^.

Presurgical evaluation begins with a non-invasive phase I assessment that combines video-scalp EEG with neuroimaging, neuropsychological assessment, and, where appropriate, genetic testing within a multidisciplinary framework^5^. When localization remains uncertain, patients undergo phase II evaluation with invasive iEEG to capture seizures and identify at the sublobar level the seizure-onset zone (SOZ), where electrographic seizure activity is first detected^7^. The clinically defined SOZ is the gold-standard ictal localization marker in current clinical practice and an important component of the broader EZ, but the two are not equivalent: the EZ may extend beyond the observed SOZ^4^. Moreover, iEEG monitoring commonly extends over several days, during which only a limited sample of a patient’s habitual seizures may be captured, whereas interictal activity is continuously available^8^. Visual assessment of the interictal EEG signals is extremely time-consuming, particularly when epileptiform activity is sparse, frequent, or widely propagated. Automated methods using continuously available interictal biomarkers could therefore complement ictal SOZ localization while reducing the amount of prolonged recording that requires manual review.

Early machine-learning approaches used handcrafted features and conventional classifiers, including logistic regression^9–14^, support vector machines^15–22^, classification trees^23–25^, and basic neural networks^26–30^, but showed limited discrimination and cross-center generalization^31^. More recently, graph neural networks (GNNs) have encoded the spatial organization of electrode implantations, improving localization over feature-based approaches^32^. However, their reliance on predefined graph connections may limit the direct modeling of long-range interactions.

Here, we introduce a topology-aware Transformer that integrates interictal neural activity and distances between recording contacts through self-attention. For each contact, self-attention allows the model to learn how much information to draw from each other contact. In GNNs that use a predefined connectivity graph, information passes directly only between connected contacts. Our model can directly capture relationships between any pair of contacts, including those far apart in the implant. Trained exclusively on public datasets, the model generalized without site-specific fine-tuning to 216 patients from five independent centers, outperforming both interictal spike-rate and clinically defined SOZ baselines. Concordance between predicted and resected tissue was strongest in patients with favorable surgical outcomes, indicating that interictal recordings alone can provide clinically meaningful spatial evidence for localizing epileptogenic tissue without requiring seizure capture.

## Results

### Multicohort development and external evaluation

We developed and externally evaluated an implant-topology-aware Transformer for contact-level localization of epileptogenic tissue from interictal iEEG. For each patient, the model integrated 30–120 min of interictal activity with electrode locations registered in MNI space to assign each recorded contact a probability of intervention (Fig. 1a). Throughout this study, we define model prediction target as tissue that was surgically removed or ablated. The fixed model was evaluated against contact-level resection labels in each patient using the area under the precision–recall curve (AUPRC) and the area under the receiver operating characteristic curve (AUROC).

**Figure 1.**
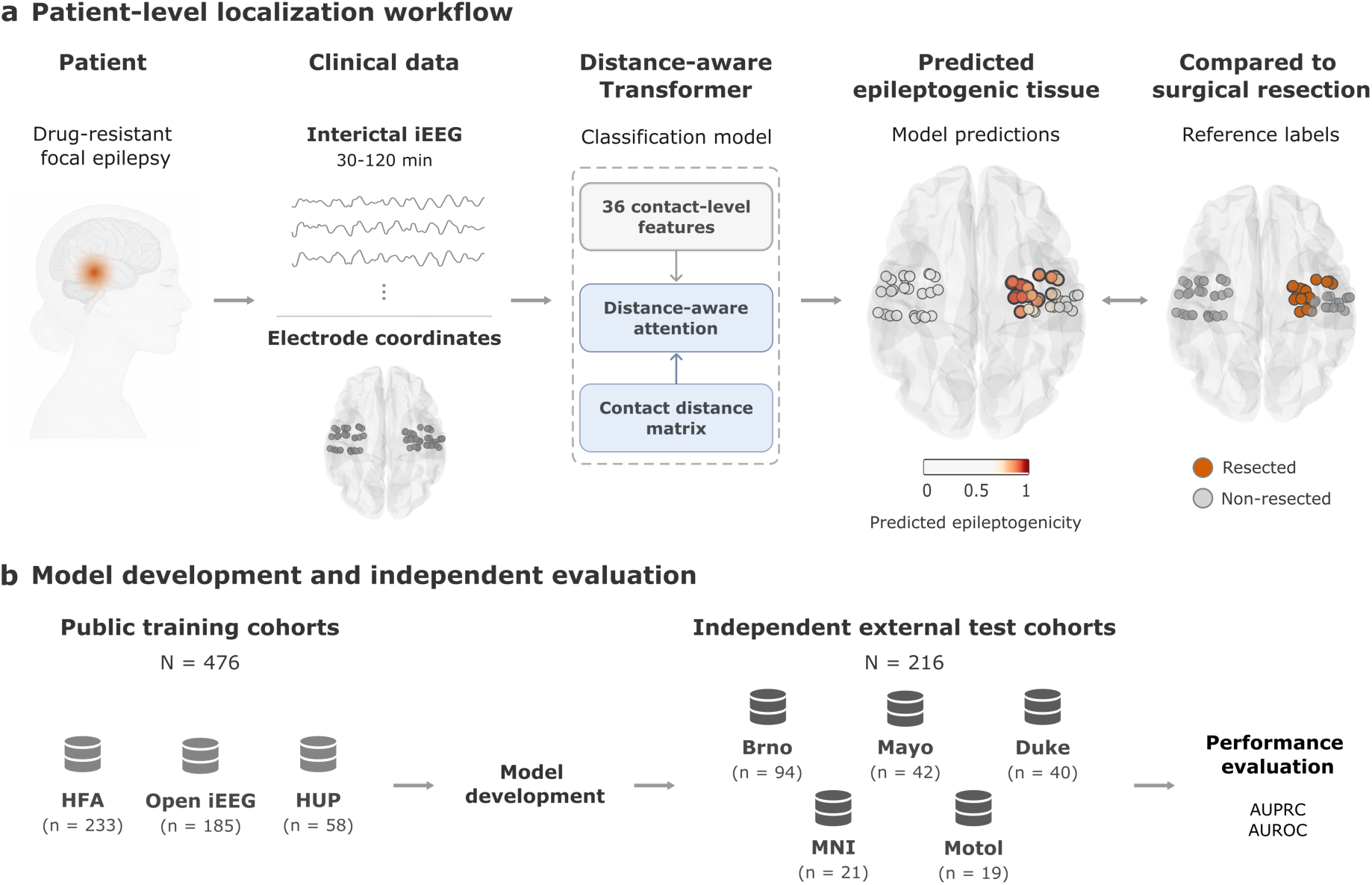
Patient-level localization workflow and independent multicenter evaluation. (a) The distance-aware transformer integrated 30–120 min of interictal intracranial EEG (iEEG) with localized electrode coordinates to generate a probability of resection for each recorded contact. Model predictions were evaluated against contact-level resection labels. Throughout this study, resected tissue encompasses tissue removed surgically or ablated. (b) Three public OpenNeuro cohorts comprising 476 patients were used in a leave-one-center-out development framework, yielding three models that formed a fixed ensemble. This ensemble was evaluated without adaptation in 216 patients from five independent external cohorts. Performance was quantified using the area under the precision–recall curve (AUPRC) and the area under the receiver operating characteristic curve (AUROC), with each patient evaluated independently.

The retrospective study included 692 patients undergoing iEEG monitoring from eight cohorts and encompassed both stereo-electroencephalography (sEEG) and electrocorticography (ECoG). Model development was restricted to 476 patients from three public OpenNeuro database cohorts^32–36^ (ds004100, ds005398, ds007118, ds007119, ds007120). Leave-one-center-out training produced three models, whose probabilities were averaged to form a fixed classification ensemble. This fixed ensemble was evaluated without further training or adaptation in 216 patients from five independent cohorts in the Czech Republic, the United States, and Canada; none contributed to model training, hyperparameter tuning, or model selection (Fig. 1b; Table 1).

**Table 1.** Model development and independent external evaluation cohorts. Three public OpenNeuro cohorts comprising 476 patients were used exclusively for model development in a leave-one-center-out framework. Five independent institutional cohorts comprising 216 patients were used exclusively for external evaluation and did not contribute to training, hyperparameter tuning or model selection. Data are presented as n or mean ± standard deviation. Surgical outcome was unavailable for 23 patients in ds005398. F, female; M, male; ECoG, electrocorticography; sEEG, stereo-electroencephalography.

| Cohort | Implant, n | Sex, F/M | Age, years | Engel I/II-IV | Total |
|---|---|---|---|---|---|
| **Model development cohorts** |  |  |  |  |  |
| iEEG comprehensive HFA model (ds007118–ds007120) | ECoG, 231;<br>sEEG, 2 | 119/114 | 13 ± 10 | 146/87 | 233 |
| Open iEEG Dataset: WSU and UCLA (ds005398) | ECoG, 167;<br>sEEG, 18 | 91/94 | 13 ± 7 | 113/49 | 185 |
| HUP iEEG Epilepsy Dataset (ds004100) | ECoG, 20;<br>sEEG, 38 | 31/27 | 34 ± 10 | 36/22 | 58 |
| Model development total | ECoG, 418;<br>sEEG, 58 | 241/235 | 16 ± 11 | 295/158 | 476 |
| **External evaluation cohorts** |  |  |  |  |  |
| Brno Epilepsy Center, Czech Republic | sEEG, 94 | 39/55 | 34.1± 10.1 | 40/54 | 94 |
| Mayo Clinic, USA | ECoG, 42 | 23/19 | 35.1±14.5 | 27/15 | 42 |
| Duke University, USA | sEEG, 40 | 22/18 | 30 ± 10.4 | 23/17 | 40 |
| Montreal Neurological Institute, Canada | sEEG, 21 | 12/9 | 31.5 ± 8.5 | 11/10 | 21 |
| Motol University Hospital, Czech Republic | sEEG, 19 | 7/12 | 22.9±11.4 | 19/0 | 19 |
| External evaluation total | ECoG, 42;<br>sEEG, 174 | 103/113 | 32.3±11.5 | 120/96 | 216 |

### Prediction–resection alignment across five centers

The fixed model ensemble was evaluated without adaptation in 216 patients from five independent institutions across three countries. Contact-level prediction–resection alignment was quantified separately for each patient and stratified by postsurgical outcome (Fig. 2).

**Figure 2.**
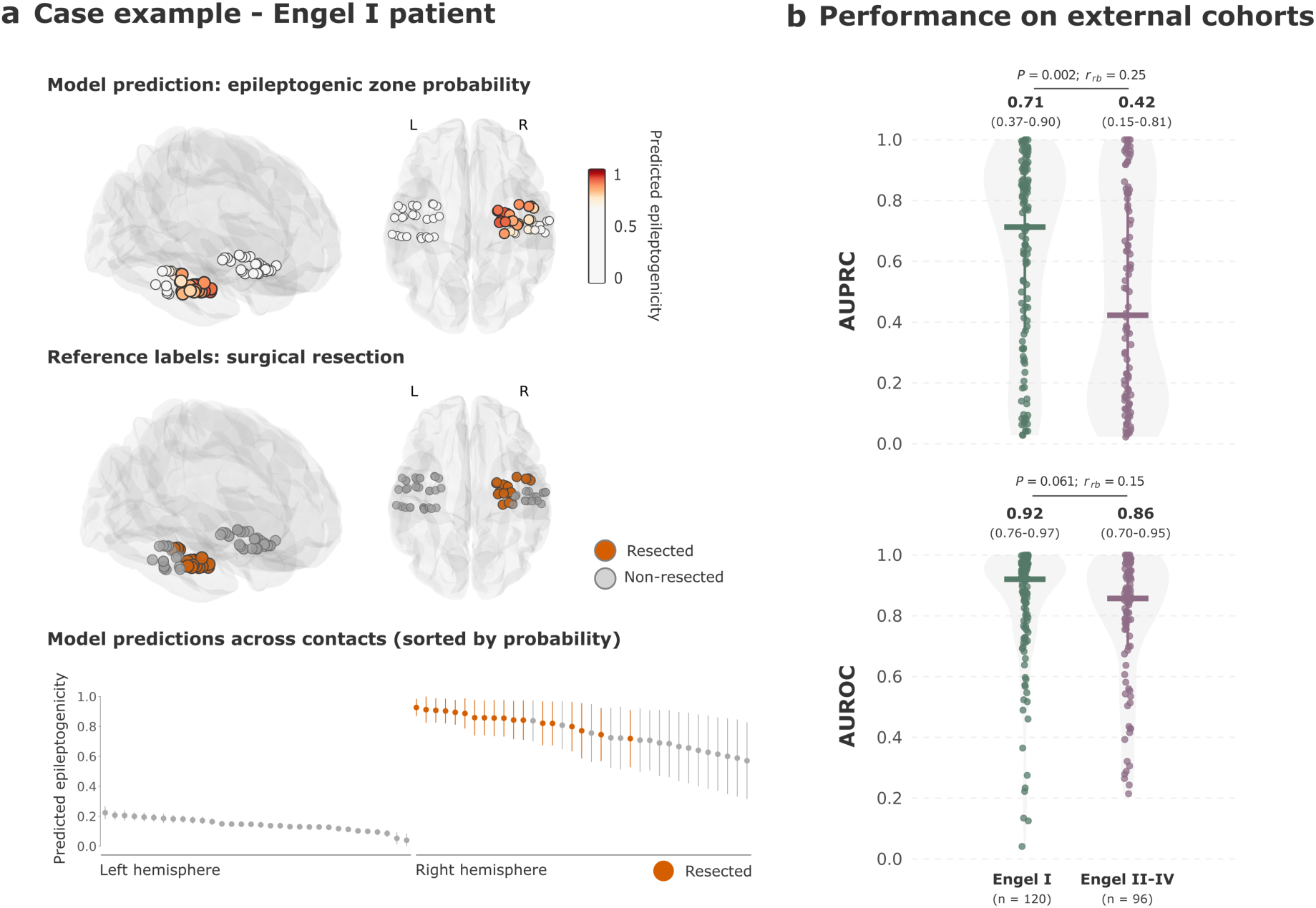
Prediction–resection alignment across independent institutional cohorts. (a) Representative contact-level predictions in a woman (30-35 years old) with a 30-year history of right temporal lobe epilepsy. MRI showed a hypotrophic right hippocampus, and SEEG localized seizure onset to the right hippocampus. Anteromesial temporal resection revealed hippocampal sclerosis (grade III), with an Engel IA outcome at 3.5 years. Each contact is colored according to its model-assigned probability, with the subsequently resected region shown in orange. High-probability contacts were concentrated within the resected region (AUPRC, 0.959; AUROC, 0.987). (b) Patient-level area under the precision–recall curve (AUPRC) and area under the receiver operating characteristic curve (AUROC), stratified by postsurgical outcome. Points represent individual patients, lines indicate medians, and group medians and interquartile ranges are shown above each distribution. Differences between patients with favorable outcomes (Engel I; n = 120) and unfavorable outcomes (Engel II–IV; n = 96) were assessed using two-sided Mann–Whitney U tests. Effect sizes are reported as rank-biserial correlations (r_rb_), with positive values indicating higher performance in Engel I. Higher values indicate closer alignment between model predictions and subsequently resected tissue.

Among patients with favorable outcomes (Engel I; n = 120), the model achieved a median AUPRC of 0.71 (IQR, 0.37–0.90) and AUROC of 0.92 (IQR, 0.76–0.97; Fig. 2b). Prediction-resection alignment was lower in patients with unfavorable outcomes (Engel II–IV; n = 96), with a median AUPRC of 0.42 (IQR, 0.15–0.81; *P* = 0.002; *rrb* = 0.25; Fig. 2b) and AUROC of 0.86 (IQR, 0.70–0.95; *P* = 0.061; *rrb* = 0.15; Fig. 2b). Median AUPRC values in both groups substantially exceeded their respective median chance reference levels of 0.149 and 0.118, defined by each patient’s proportion of resected contacts (Supplementary Table 1).

Institution-specific analyses showed broadly consistent prediction–resection alignment across the five external cohorts (Supplementary Figs. 1 and 2). Among patients with Engel I outcomes, median AUROC ranged from 0.86 to 0.97 and median AUPRC from 0.63 to 0.87 across institutions. Performance was numerically higher in Engel I than in Engel II–IV patients across all four cohorts that included both outcome groups (per institution results are reported in Supplementary Table 1).

### Prediction–resection alignment by epileptogenic zone location

To assess whether prediction–resection alignment depended on the anatomical extent of the resection, patients were stratified into temporal (n = 78), temporal-plus (n = 48), and extratemporal (n = 90) groups (Fig. 3c). Resections were classified according to position of resected regions based on MNI atlas. The temporal-plus indicates that resected contacts were in both temporal and extratemporal regions. In patients with temporal resections, alignment was high irrespective of postsurgical outcome, with a median AUPRC of 0.77 in Engel I and 0.67 in Engel II–IV patients (P = 0.471). In contrast, AUPRC was lower in patients with unfavorable outcomes in both temporal-plus (0.68 vs 0.26; P = 0.020) and extratemporal resections (0.66 vs 0.23; P = 0.039). The outcome-related difference observed in the full cohort therefore arose predominantly from patients with temporal-plus and extratemporal resections, in whom the highest-ranked contacts more often lay outside the resection when outcomes were unfavorable.

**Figure 3.**
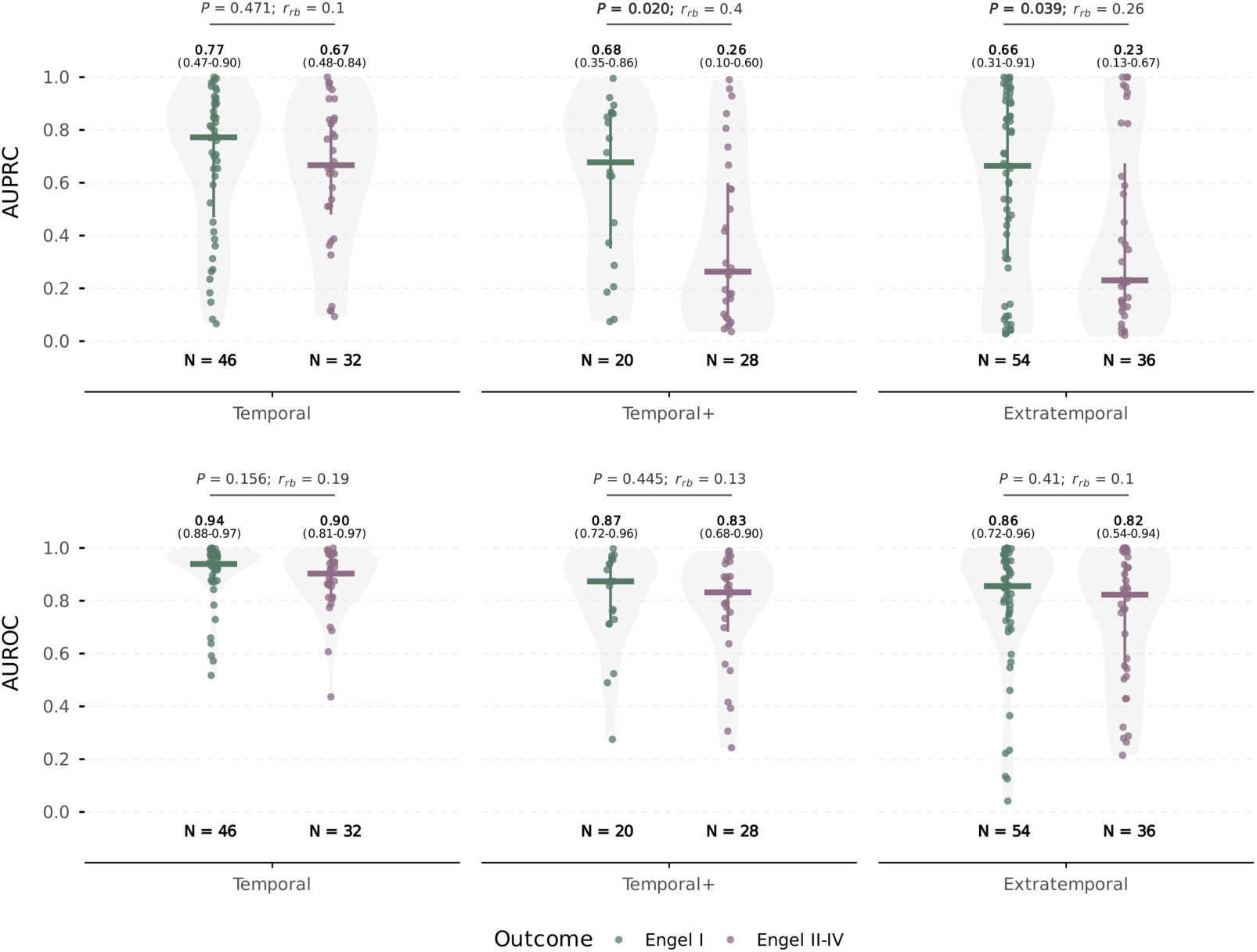
Prediction–resection alignment stratified by resection extent. Patient-level AUPRC (top) and AUROC (bottom) for temporal (n = 78), temporal-plus (n = 48; resected contacts in both temporal and extratemporal regions), and extratemporal (n = 90) resections, stratified by outcome (Engel I, green; Engel II–IV, purple). Points represent individual patients; lines indicate medians and interquartile ranges, which are shown above each distribution. Outcome groups were compared using two-sided Mann– Whitney U tests. Effect sizes are rank-biserial correlations, with positive values indicating higher performance in Engel I. In patients with temporal resections, alignment was high irrespective of postsurgical outcome, with a median AUPRC of 0.77 in Engel I and 0.67 in Engel II–IV patients (P = 0.471). In contrast, AUPRC was lower in patients with unfavorable outcomes in both temporal-plus (0.68 vs 0.26; P = 0.020) and extratemporal resections (0.66 vs 0.23; P = 0.039). AUROC did not differ significantly between outcome groups in any subgroup (median 0.82–0.94). The outcome-related difference observed in the full cohort therefore arose predominantly from patients with temporal-plus and extratemporal resections, in whom the highest-ranked model predictions often lay outside the resection when outcomes were unfavorable.

### Thresholding trades sensitivity for spatial precision

In patients with a favorable outcome (Engel I), we examined operating characteristics obtained by thresholding model probabilities from 0.75 to 0.90 and spatially expanding the selected contact sets by 3–12 mm (Table 2). Increasing the spatial neighborhood radius increased sensitivity but reduced specificity and positive predictive value (PPV), whereas increasing the probability threshold reduced sensitivity while increasing specificity and PPV. Median sensitivity reached 100% at a probability threshold of 0.75 and a 12-mm radius. Conversely, the highest median specificity (94%) and PPV (62%) were observed at a threshold of 0.90 and a 3-mm radius.

**Table 2.** Threshold-dependent operating characteristics in patients with favorable postsurgical outcomes. Continuous model probabilities were converted into binary intervention suggestions by selecting contacts exceeding the specified probability threshold and expanding the selection to contacts within the specified resection radius. Values are group medians. SEN, sensitivity; SPE, specificity; PPV, positive predictive value.

| SEN/SPE/PPV (%) | Probability Threshold |  |  |  |
| Resection Radius | 0.75 | 0.80 | 0.85 | 0.90 |
|---|---|---|---|---|
| 3 mm | 83 / 78 / 43 | 72 / 84 / 45 | 58 / 90 / 52 | 40 / 94 / 62 |
| 6 mm | 88 / 75 / 40 | 78 / 82 / 42 | 65 / 88 / 47 | 48 / 93 / 57 |
| 9 mm | 92 / 73 / 35 | 81 / 80 / 40 | 72 / 85 / 48 | 53 / 91 / 56 |
| 12 mm | 100 / 68 / 34 | 87 / 76 / 36 | 82 / 83 / 45 | 63 / 88 / 53 |

### Comparison with interictal and clinical baselines

We compared model prediction–resection alignment with interictal spike rate, the clinically annotated SOZ, and the SOZ expanded by a 10-mm spatial margin (SOZ+10 mm). The expanded SOZ baseline was included to reduce sensitivity to variability in the number and spatial extent of contacts annotated as SOZ across patients. These comparisons were limited to patients with Engel I outcomes (Fig. 4a).

**Figure 4.**
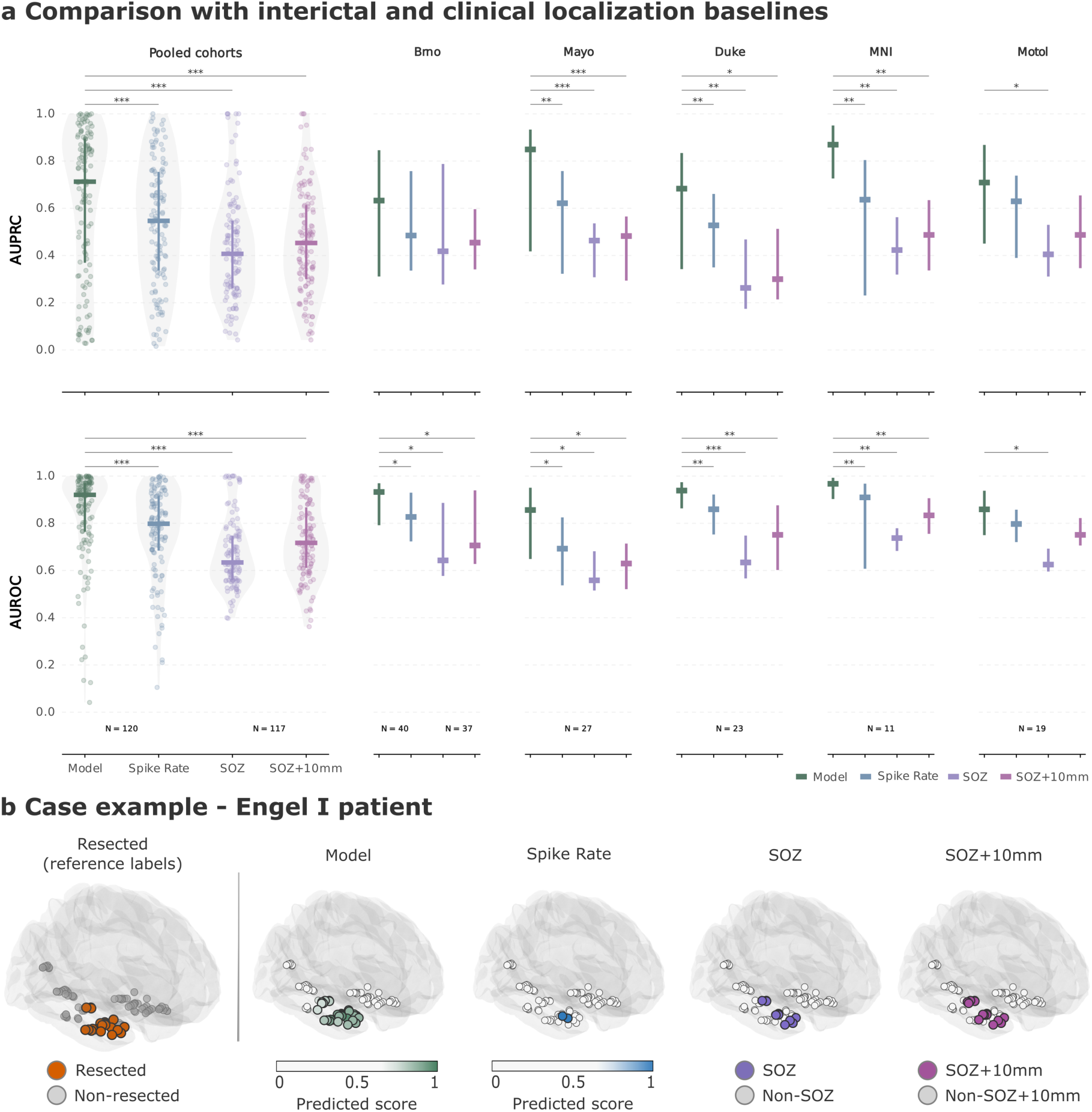
Comparison with interictal and clinical localization baselines. (a) Patient-level area under the precision–recall curve (AUPRC; top) and area under the receiver operating characteristic curve (AUROC; bottom) for the distance-aware Transformer ensemble, interictal spike rate, the clinically annotated seizure-onset zone (SOZ), and the SOZ expanded by a 10-mm Euclidean margin (SOZ+10 mm). Analyses were restricted to patients with favorable postsurgical outcomes (Engel I). SOZ was not identified for 3 patients from the Brno cohort. Pooled distributions are shown in the left column; institution-specific medians and interquartile ranges are shown in the remaining columns. Points represent individual patients, horizontal bars indicate medians, and vertical lines indicate interquartile ranges. Comparisons between the model and each baseline were performed using two-sided paired Wilcoxon signed-rank tests under the prespecified alternative hypothesis that model performance was higher. P values were adjusted using the Benjamini–Hochberg false discovery rate (FDR) procedure. ∗ P_FDR_ < 0.05, ∗∗ P_FDR_ < 0.01 and ∗∗∗ P_FDR_ < 0.001; unmarked comparisons were not significant. (b) Representative contact-level localization in a woman (35-40 years old) with a 27-year history of right temporal lobe epilepsy. MRI showed bilateral hippocampal atrophy and malrotation, whereas SEEG localized seizure onset to the right hippocampus. Right anteromesial temporal resection yielded negative histopathological findings, with an Engel IB outcome at 10 years. Model performance was AUPRC 0.902 and AUROC 0.969. Resected contacts are shown in orange in the reference map. Model predictions and spike rates are displayed as continuous scores; SOZ and SOZ+10 mm are binary contact classifications.

Across the pooled external cohorts (Supplementary Table 2), the model achieved a median AUPRC of 0.71, compared with 0.55 for spike rate and 0.41 for SOZ, and 0.45 for SOZ+10mm. Median AUROC was 0.92 for the model, compared with 0.80 for spike rate, 0.63 for SOZ, and 0.72 for SOZ+10 mm. Pooled results of the proposed model outperformed all three baselines for both AUPRC and AUROC in paired analyses (all false-discovery-rate-adjusted values, *P*_FDR_ < 0.001). In the representative Engel I case, high model scores extended across the resected contact cluster more consistently than the spike-rate and SOZ-based localizations (Fig. 4b).

Institution-specific analyses confirmed that the pooled advantage was not driven by a single cohort (Fig. 4a and Supplementary Table 2). At Mayo, Duke, and MNI, the model significantly outperformed all three baselines for both AUPRC and AUROC. In Brno, it significantly outperformed all baselines on AUROC. At Motol, significant improvements were observed primarily relative to the SOZ-based baseline.

### Peak predictions are displaced in unfavorable outcomes

To characterize prediction–resection spatial alignment, we measured the Euclidean distance between each patient’s highest-confidence model prediction (*p*_max_) and the nearest resected contact. In the pooled external cohorts, *p*_max_ corresponded to a resected contact in approximately 68.3% of Engel I patients, compared with 47.9% of Engel II–IV patients. Within 10 mm of a resected contact, the corresponding proportions were 72.5% and 57.3%, increasing to 83.3% and 69.8% within 20 mm (Fig. 5). Overall, distances were smaller in patients with Engel I outcomes (median, 0 mm; IQR, 0–11.2 mm) than in those with Engel II–IV outcomes (median, 3.72 mm; IQR, 0–24.2 mm; two-sided Mann–Whitney *U*-test, *U* = 4,612.5, *P =* 0.005, *rrb =* 0.20). The cumulative distance distribution was shifted toward greater distances in Engel II–IV patients in the pooled analysis, indicating that their highest-confidence predictions were more frequently located outside the resected tissue. Similar separation was apparent in Brno and MNI, whereas the distributions overlapped more extensively in Mayo and Duke; Motol included only Engel I patients (Supplementary Fig. 3).

**Figure 5.**
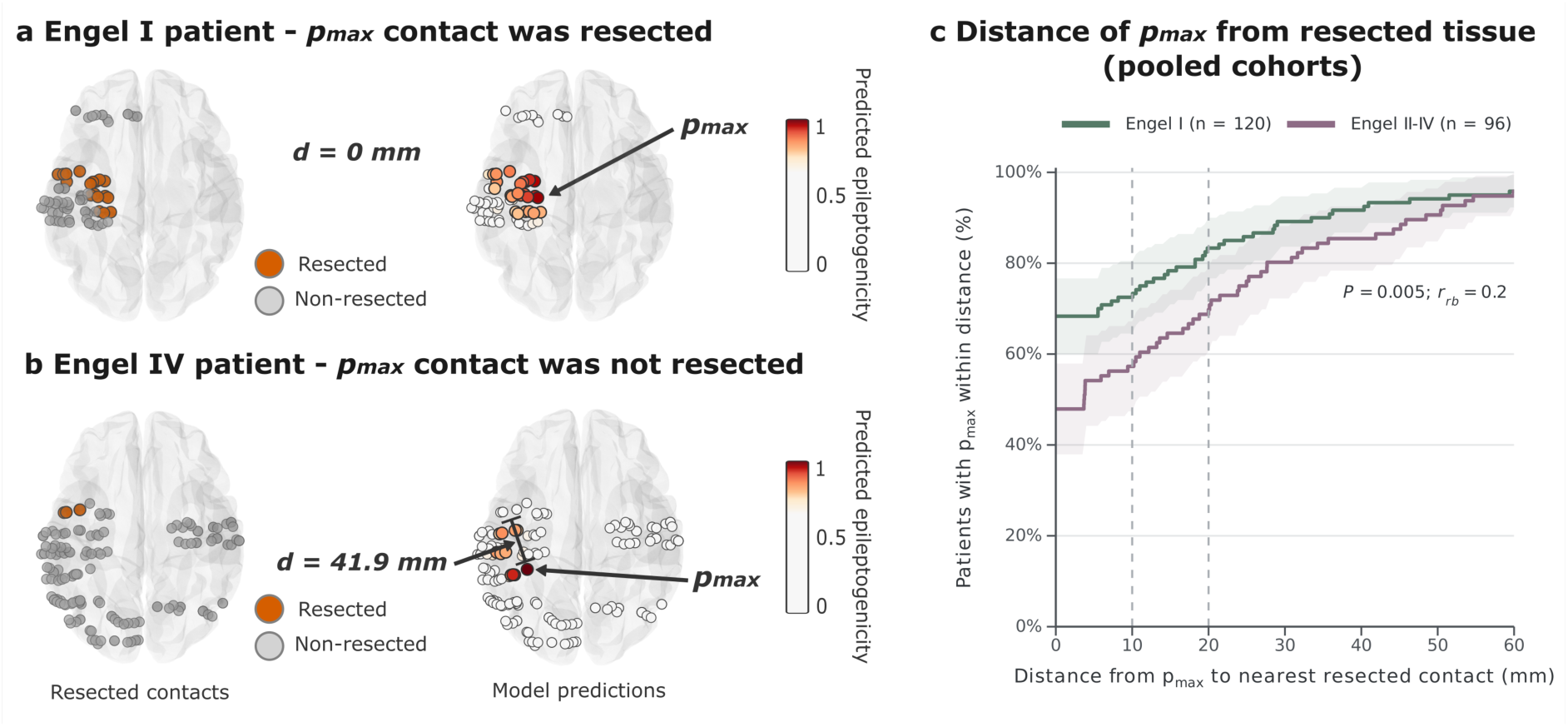
Spatial displacement of peak model predictions relative to resected tissue. (a,b) Representative patients with Engel I (a) and Engel IV (b) postsurgical outcomes. For each patient, the left figure shows resected (orange) and non-resected (grey) electrode contacts, and the right rendering shows contact-level predicted epileptogenicity. The contact receiving the highest model probability (p_max_) and its Euclidean distance (d) to the nearest resected contact are indicated. In the Engel I example, the p_max_ contact was resected (d = 0 mm); in the Engel IV example, it was located 41.9 mm from the nearest resected contact. (c) Pooled empirical cumulative distributions of d across the external cohorts, stratified by postsurgical outcome. At each distance, the y-axis indicates the proportion of patients whose p_max_ was located within that distance of a resected contact. Shaded bands indicate 95% confidence intervals. Vertical dashed lines mark 10 and 20 mm. Distances were significantly smaller in Engel I than in Engel II–IV patients (two-sided Mann–Whitney U-test; rank-biserial correlation).

## Discussion

Computational approaches to localizing the EZ from iEEG have largely analyzed contacts in isolation, for example, through rates of interictal epileptiform discharges or high-frequency oscillations, or characterized functional connectivity without explicitly accounting for implant topology and anatomical context. Here, we present an implant topology-aware Transformer that integrates interictal activity with the spatial organization of implanted contacts to rank candidate treatment sites. Trained exclusively on public datasets, the model assigned higher probabilities to tissue subsequently treated in an external cohort of 216 patients from five independent centers, without site-specific adaptation. Agreement was greatest in patients who achieved seizure freedom. The preservation of performance across datasets demonstrates the feasibility of transferring topology-aware interictal iEEG analysis across populations, centers, and implantation modalities. This evaluation establishes generalizability beyond the training data as a proof of principle; future models trained on pooled, diverse cohorts may further improve performance. The results support topology-aware attention as a promising framework for interictal iEEG analysis.

A key methodological distinction is how the model represents relationships between implanted contacts. Approaches based on individual-channel features, such as spike rates, can identify locally abnormal activity but do not explicitly account for the spatial context in which each contact was recorded. Functional connectivity methods capture relationships between signals, but these relationships alone do not necessarily reflect the physical organization of the implant or the anatomical distances between sampled sites. GNNs incorporate electrode relationships into the learning process, although message passing commonly relies on predefined neighborhoods and may progressively smooth differences between contacts as information propagates through the graph. The proposed Transformer model uses an attention mechanism to capture relationships across the implanted contacts, integrating spatial information with interictal features. Rather than assigning each contact an isolated label, this enables each contact to be interpreted in the context of the patient-specific implantation.

Incorporating anatomical coordinates provides additional context by allowing the model to represent both the relative arrangement of contacts and their absolute locations in the brain. This may help identify spatial patterns in interictal activity that are not captured by electrophysiological features alone and may support transfer across different implantation configurations. At the same time, absolute positional encoding introduces the possibility that the model learns regional patterns of clinical practice or treatment selection. If certain brain regions are more frequently sampled or resected in the development cohorts, anatomical location could become associated with treatment probability independently of its electrophysiological relevance, which we described in our previous work with GNNs^31^. This concern is particularly important when cohorts differ in the composition of temporal and extratemporal epilepsy. Anatomical encoding is thus a potentially valuable source of information, but its contribution must be assessed alongside the risk of regional bias.

The external clinical evaluation provides evidence that the model captures information relevant to treatment localization beyond clinically defined SOZ baselines. Interictal activity may therefore provide complementary information that is available outside individual seizures. The model’s closer alignment with treated tissue in patients who achieved seizure freedom is consistent with the possibility that successful treatment more often encompassed tissue carrying a prominent interictal signature. Conversely, weaker alignment in patients with persistent seizures could reflect a more distributed epileptogenic network, incomplete sampling or treatment, or other differences in disease and clinical management. These explanations remain hypotheses and cannot be distinguished by outcome associations alone. Moreover, the location of a single highest-ranked contact provides only a partial view of a patient-level ranking and does not fully characterize the spatial distribution of predicted probabilities across the implant.

A principal strength of this study is the scale and independence of the evaluation. Previous automated localization studies have generally included smaller cohorts and relied on internal validation or data from a limited number of centers. Here, model development was confined to public datasets, and the fixed ensemble was applied without retraining or recalibration to 216 patients from five external institutions. The development cohorts were predominantly recorded with ECoG, whereas the external cohorts were predominantly recorded with sEEG. Alignment was nevertheless retained across this shift in population and implantation modality, which reduces the likelihood that the reported performance reflects institution-specific acquisition practices or adaptation to the evaluation cohorts.

The resulting performance estimates compare favorably with those reported for earlier automated multifeatured localization approaches (Table 3). Comparisons include studies using partially overlapping datasets and studies using independent datasets identified through a systematic review by Hrtonova et al.^33^, with selection criteria detailed in the Methods. A direct ranking is not possible since studies differ in datasets, reference labels, patient selection, and outcome definitions. The main contribution is therefore not a numerical improvement alone, but the demonstration that an implant topology-aware model trained exclusively on public data can maintain prediction–resection alignment across several independent clinical cohorts without retraining.

**Table 3.** Comparison of published methods for localizing epileptogenic tissue from intracranial EEG. Studies are grouped by whether their datasets partially overlap with those used in the present study or are independent. Target denotes the contacts labeled positive for performance evaluation: Resected, contacts within the surgical resection; Resected SOZ, SOZ contacts within the resection; SOZ, clinician-defined seizure-onset zone contacts; SOZ*, an extended clinical target comprising the SOZ and early propagation contacts (Gunnarsdottir et al.) or the SOZ and exclusively irritative zone (von Ellenrieder et al.). Outcomes are reported as Engel I, Engel II–IV, or all included patients irrespective of outcome (All). Performance values are reported as summarized in each publication; differences in targets, cohorts, and summary statistics limit direct comparisons. Chance AUPRC denotes the prevalence of target contacts, providing the random-classifier baseline; the corresponding AUROC baseline is 0.5. n, number of patients; AUPRC, area under the precision–recall curve; AUROC, area under the receiver operating characteristic curve; GNN, graph neural network; LR, logistic regression; SVM, support vector machine; —, not reported.

| **Method** | **Target** | **Engel outcome** | ***n*** | **AUPRC** | **Chance AUPRC** | **AUROC** |
|---|---|---|---|---|---|---|
| **Distance-aware Transformer (present study)** | Resected | I | 120 | 0.71 | 0.15 | 0.92 |
|  |  | II–IV | 96 | 0.42 | 0.12 | 0.86 |
| **Partially overlapping datasets** |  |  |  |  |  |  |
| GNN (Nejedly *et al.*)<sup>32</sup> | Resected | I | 31 | 0.68 | 0.10 | 0.91 |
|  |  | II–IV | 49 | 0.38 | 0.08 | 0.83 |
| LR (Klimes *et al.*)<sup>34</sup> | Resected | I | 18 | 0.61 | 0.10 | 0.83 |
| SVM (Cimbalnik *et al.*)<sup>16</sup> | Resected | I | 9 | — | — | 0.84 |
|  |  | II–IV | 7 | — | — | 0.78 |
| SVM (Chybowski *et al.*)<sup>35</sup> | Resected SOZ | I | 25 | 0.49 | 0.06 | — |
| **Independent datasets** |  |  |  |  |  |  |
| Random Forest<br>(Bernabei *et al.*)<sup>23</sup> | Resected | I | 38 | 0.25 | — | — |
|  |  | II–IV | 22 | 0.14 | — | — |
|  |  | All | 60 | — | — | 0.77 |
| LR (Jeong *et al.*)<sup>36</sup> | Resected SOZ | I | 17 | — | — | 0.72 |
| LR (Conrad *et al.*)<sup>37</sup> | SOZ | I | 24 | — | — | 0.82 |
|  |  | All | 95 | — | — | 0.83 |
| SVM (Varatharajah *et al.*)<sup>20</sup> | SOZ | All | 82 | — | — | 0.79 |
| LR (Gunnarsdottir *et al.*)<sup>10</sup> | SOZ* | I | 28 | — | — | 0.77 |
| LR (von Ellenrieder *et al.*)<sup>38</sup> | SOZ* | All | 45 | — | — | 0.83 |

### Limitations

Multiple limitations should be considered. First, resection or ablation was used as an operational target, but treated tissue is an imperfect proxy for the EZ and reflects clinical judgment, surgical accessibility, and institutional practice. Second, the development and external cohorts differed in age, recording modality, and other aspects of data acquisition and clinical workflow. Although successful external validation across these differences supports generalizability, it does not rule out residual cohort effects. Third, iEEG samples only the regions selected for implantation; tissue outside the sampled network cannot be directly ranked, and the model’s predictions depend on the contacts available for analysis. Fourth, anatomical coordinates may encode regional treatment patterns, and differences in the representation of temporal and extratemporal cases may affect performance. Finally, the interictal segments analyzed were selected two hours away from seizures, and seizure timing was available. The present findings, therefore, do not establish that seizure capture can be omitted or that monitoring duration can safely be shortened. Further validation across broader cohorts, implantation strategies, and prospective clinical workflows is needed to determine the model’s role in epilepsy surgery planning.

### Conclusion

The model provides a scalable and objective framework for extracting localization evidence from interictal iEEG, potentially transforming how invasive recordings are used in presurgical evaluation. By leveraging minutes of interictal data rather than captured seizures, this approach could reduce reliance on prolonged hospitalizations and help inform treatment decisions even when seizures are infrequent or not recorded. More broadly, our findings challenge the notion that seizure capture is required to identify epileptogenic tissue and suggest that latent spatial biomarkers of epileptogenicity are present in interictal brain activity. Prospective studies are required to determine whether incorporating these predictions into surgical planning improves clinical decisions and patient outcomes, and whether they can reduce reliance on prolonged seizure monitoring.

## Methods

### Study design and reporting

We conducted a retrospective, multi-cohort study to develop and externally evaluate a contact-level model for localizing tissue subsequently treated during epilepsy surgery. Model development was restricted to three publicly available cohorts, whereas external evaluation used five independent institutional cohorts that were withheld from training, hyperparameter selection, model selection, and all intermediate evaluations. The unit of model prediction was the iEEG contact, and performance metrics were calculated separately for each patient.

### Setting and data sources

Model development and internal cross-validation used three OpenNeuro resources: the iEEG Comprehensive HFA Model datasets^39–41^ (ds007118–ds007120; n = 233), the Open iEEG Dataset^42^ from Washington University and the University of California, Los Angeles (ds005398; n = 185), and the HUP iEEG Epilepsy Dataset^43^ (ds004100; n = 58). Together, these datasets comprised data from 476 patients across three institutions.

External evaluation used retrospective cohorts from the Brno Epilepsy Center, Czech Republic (n = 94); Mayo Clinic, Rochester, USA (n = 42); Duke University, USA (n = 40); the Montreal Neurological Institute, Canada (n = 21); and Motol University Hospital, Prague, Czech Republic (n = 19), giving 216 externally evaluated patients. These cohorts included sEEG and ECoG recordings collected during routine presurgical evaluation at tertiary epilepsy centers.

### Ethics

The study was approved by the Brno Epilepsy Center Research Ethics Committee (EK-FNUSA-11/2024), Mayo Clinic Institutional Review Board, Duke University, the MNI Ethics Review Board, and Motol University Hospital. All participants in the external institutional cohorts provided written informed consent in accordance with the Declaration of Helsinki.

Publicly available, de-identified datasets were obtained from OpenNeuro and reused under the Creative Commons CC0 public-domain dedication. Under the OpenNeuro data-sharing terms, the original dataset contributors confirmed that they held the necessary ethics permissions to share the data publicly and that the deposited datasets contained no identifiable protected health information.

### Participants, eligibility, and follow-up

Patients were eligible if they (1) underwent presurgical invasive monitoring with sEEG or ECoG; (2) subsequently underwent resection, ablation or a combined intervention; (3) had preoperative MRI and post-implantation CT or MRI suitable for electrode localization and registration to Montreal Neurological Institute (MNI) space; (4) had sufficient interictal iEEG for feature extraction; and (5) had at least 12 months of postoperative follow-up with outcome classified using the Engel scale or the International League Against Epilepsy (ILAE) outcome scale. Patients were excluded if required imaging, electrode coordinates, resection labels or analyzable iEEG were unavailable or failed quality control. The most recent eligible postoperative assessment was used to assign the outcome. Engel class I was defined as a favorable outcome, and Engel classes II–IV as unfavorable outcomes.

### Variables and clinical annotations

The supervised target was a binary contact-level indicator of whether the tissue sampled by a contact was subsequently treated. Throughout this study, the term “resected tissue” was used operationally to encompass all treated tissue, including tissue removed by surgical resection and tissue intervened by ablation, either by sEEG-guided radiofrequency thermocoagulation (RFTC) or laser ablation. Resected contacts were identified based on pre- and post-surgical MRI. Contacts classified as lying within the resected volume were assigned a positive label; all other analyzed contacts were assigned a negative label.

The principal model predictors comprised interictal event rates, event-propagation measures, spectral features, gray/white matter tissue-probability values, and implantation-topology descriptors. Postsurgical outcome group and institution were used as stratification variables. Clinical baselines were interictal spike rate, the clinically annotated SOZ, and the SOZ expanded to include contacts within a 10-mm Euclidean distance of an SOZ contact (SOZ+10 mm). SOZ annotations were established at each participating institution as part of routine clinical evaluation, before application of the model and without knowledge of its outputs. Definitions and annotation procedures were not retrospectively harmonized across institutions, thereby preserving the clinical heterogeneity of the independent datasets.

### Electrode localization and spatial measurements

Electrode contacts were localized from post-implantation CT or MRI, coregistered to pre-operative MRI, and transformed to MNI space. Pairwise Euclidean distances between all contacts were calculated in millimeters from their MNI coordinates. Grey-matter, white-matter and cerebrospinal-fluid probabilities were sampled at each coordinate from a tissue-probability atlas.

### iEEG selection and preprocessing

For each iEEG recording, 30–120 min of interictal data were manually selected from periods at least 2 h distant from any seizure and free of ictal or immediate postictal activity. Recordings were acquired in a referential montage at a sampling frequency of ≥500 Hz during sleep or resting wakefulness, depending on data availability. Recordings were divided into non-overlapping 10-min windows, and features were calculated independently for each contact and window. For model development, segments of at least 5 min were accepted when a complete 10-minute segment was not available. Center-specific acquisition and initial processing procedures are summarized in Supplementary Table 3. All recordings subsequently underwent the same feature-extraction pipeline.

### Feature extraction

Each contact was represented by 36 scalar features comprising two interictal event-rate features, six network-propagation features, 15 spectral features, three tissue-probability features, three stereotactic coordinates, and seven implantation-geometry features (Table 4). Features were first computed separately for each recording segment, then averaged across segments to obtain a single value per feature for each contact and recording. All transformations were calculated independently within each patient segment rather than using scaling parameters estimated from the training cohort.

**Table 4.** Summary of the 36 contact-level input features and their processing. Features were calculated separately for each recording segment and then averaged across segments to obtain a single value per feature for each contact and recording. Event rates were expressed as events per minute and transformed as log(1+x), where x is the event rate. Maximum scaling and percentile ranking were performed across contacts within each segment. CSF, cerebrospinal fluid; HITS, Hyperlink-Induced Topic Search; ICBM, International Consortium for Brain Mapping; MNI, Montreal Neurological Institute; PSD, power spectral density; SD, standard deviation. Frequencies are reported in hertz (Hz), temporal windows in milliseconds (ms), and distances in millimeters (mm).

| Category | Features | Processing |
|---|---|---|
| Event rates | Spike rate; ripple rate | Events/min; log(1+x) |
| Event propagation | Spike hub and authority; ripple hub and authority; spike-to-ripple hub and authority | 100-ms window; five nearest targets; within-segment percentile rank |
| Aperiodic spectrum | Spectral slope; spectral offset | Welch PSD; Huber fit over 1–40 Hz |
| Absolute power | 0.1–1, 1–4 and 30–80 Hz | Within-segment maximum scaling |
| Phase–amplitude coupling | Delta–gamma; delta–ripple | Tort modulation index; 18 phase bins; maximum scaling |
| Relative power | 0.1–1, 1–4, 4–8, 8–12, 12–30, 30–80, 45–65 and 80–250 Hz | Fraction of total spectral power |
| Tissue probability | Grey matter; white matter; CSF | ICBM152 nonlinear symmetric 2009a atlas |
| MNI position | Right–left; anterior–posterior; inferior–superior coordinates | No normalization |
| Implant geometry | Mean, SD, minimum and maximum distance; neighbor counts within 10, 20 and 30 mm | Euclidean distance in MNI space; no normalization |

Interictal epileptiform spikes were detected using the automated signal-envelope distribution method described by Janca et al.^41^ Ripple events were detected by Hilbert detector applying the corresponding band-event detector to signals filtered between 80 and 250 Hz. For each contact, spike and ripple rates were calculated as the number of detected events divided by the segment duration in minutes. Rates were transformed as log(1 + *x*), where *x* denotes the event rate in events per minute, thereby retaining contacts with zero detected events while reducing the influence of very high rates.

Propagation features were derived from three directed event networks: spike-to-spike, ripple-to-ripple and spike-to-ripple networks. For each source event, potential target events occurring within the subsequent 100 ms were identified, and edges were constructed using up to five temporally nearest target events. Edge weights represented the accumulated number of source– target associations across the segment. Spike-to-ripple networks used spikes as source events and ripples as target events. The Hyperlink-Induced Topic Search algorithm^44^ was applied separately to each adjacency matrix to obtain one hub and one authority score per contact, yielding six features. Hub scores characterize contacts whose events tended to precede events at other contacts, whereas authority scores characterize contacts whose events tended to follow activity elsewhere. The resulting scores were converted to within-segment percentile ranks by dividing each contact’s rank by the number of contacts in that segment. This procedure was related to the network-based approach described by Nejedly et al.^32^

Spectral features comprised the slope and offset of the aperiodic spectrum, three absolute band-power measures, two phase–amplitude coupling measures, and eight relative band-power measures. Power spectral density was estimated using Welch’s method^45^. The aperiodic component was estimated over 1–40 Hz by fitting a Huber robust linear regression to *log*_10_-transformed frequency and power values; the fitted slope and intercept were used without further normalization. Absolute power was calculated in the low-delta (0.1–1 Hz), delta (1–4 Hz) and gamma (30–80 Hz) bands. Frequencies between 45 and 55 Hz were excluded from the gamma-power calculation to reduce line-noise contamination. A 10-s Welch window was used for low-delta power, whereas 1-s windows were used for delta and gamma power. Each absolute-power feature was divided by its maximum across contacts within the same segment; segments with a maximum value of 1 × 10^−8^ or lower were assigned zero for that feature.

Phase–amplitude coupling was quantified between delta phase (1–4 Hz) and the amplitude of either gamma activity (30–80 Hz) or ripple activity (80–250 Hz). Band-limited phase and amplitude were obtained using Butterworth filtering followed by the Hilbert transform. Amplitude values were grouped into 18 phase bins, and coupling strength was quantified using the Tort modulation index^46^, 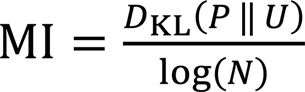, where *P* is the observed phase-binned amplitude distribution and *U* is the uniform distribution. Each coupling feature was divided by its maximum across contacts within the same segment.

Relative spectral power was calculated in eight bands: 0.1–1, 1–4, 4–8, 8–12, 12–30, 30–80, 45–65, and 80–250 Hz. The power spectral density was normalized to a total power of 1 before summing within each frequency band. A 10-s Welch window was used for the 0.1–1-Hz band and a 1-s window for the remaining bands. Because these features were already expressed as fractions of total power, no additional normalization was applied.

Anatomical context was represented by the probabilities that each contact was located in grey matter, white matter, or cerebrospinal fluid. Probabilities were obtained from the ICBM152 nonlinear symmetric 2009a tissue-probability maps^47^ by transforming each contact’s MNI coordinate to voxel space and sampling the nearest atlas voxel without interpolation. Coordinates outside the atlas bounds were assigned a probability of zero. The contact’s right– left, anterior–posterior, and inferior–superior MNI coordinates were also included as three unnormalized features.

Implantation geometry was summarized using the full matrix of pairwise Euclidean distances between contacts in MNI space. Furthermore, for each contact, seven scalar descriptors were calculated: the mean, standard deviation, minimum, and maximum distance to all contacts, together with the number of contacts located within 10, 20, and 30 mm. These features were not normalized. The full *N* × *N* distance matrix, expressed in millimeters, was supplied separately to the distance-aware attention mechanism and was not included among the 36 per-contact features.

### Distance-aware Transformer

The model framed localization as supervised binary node classification over the implanted electrode contacts (Fig. 6) forming computational graph, where nodes are described by node features and edges are relative Euclidean distances. The 36 input features for each contact were linearly projected into a uniform 32-dimensional embedding space. In general, conventional transformer architecture requires absolute or relative positional encodings for node description. Here we deliberately use MNI coordinates (absolute) and neighborhood features (relative) as node positional encodings. At the same time, we use attention bias derived from edge features (for this purpose Euclidean distance). In general, this can be used by any electrode-electrode interaction such as functional connectivity. Implantations were zero-padded or truncated to a maximum of 256 contacts. A Boolean mask identified valid contacts and was applied to the attention logits and loss, preventing padded positions from being attended to or contributing to model optimization.

**Figure 6.**
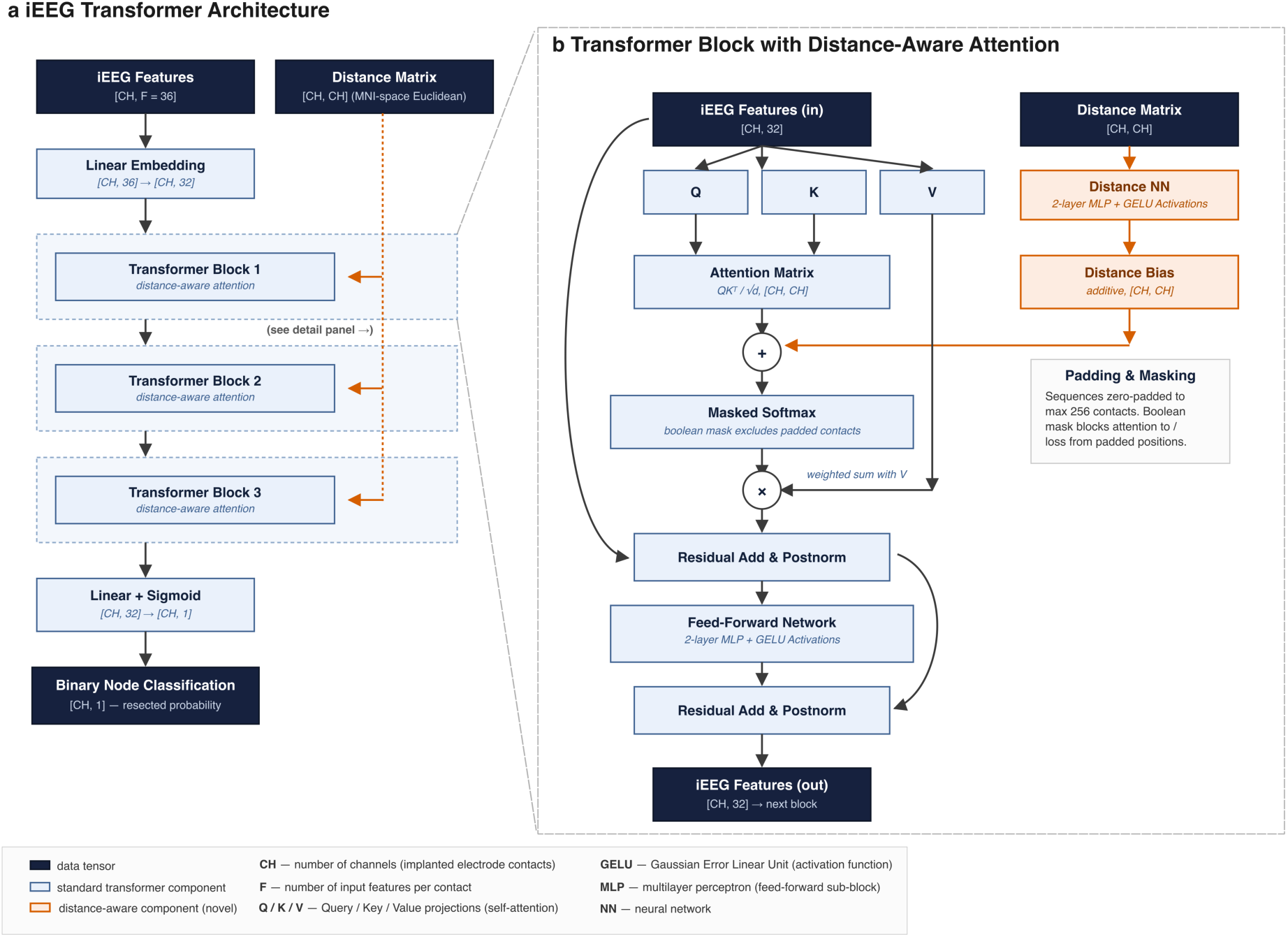
Distance-Aware Transformer architecture. (a) Overall model architecture. Electrode-contact features and the pairwise inter-contact distance matrix (MNI-space Euclidean distances) are input in parallel; features are linearly embedded and passed through three stacked Transformer blocks with distance-aware attention (detail in panel b), followed by a linear classifier producing a per-contact probability of resection. (b) Detail of a single Transformer block. Standard query/key/value self-attention is supplemented with a learned additive bias computed from the distance matrix by a dedicated Distance Neural Network (orange), summed with the attention logits prior to a masked softmax that excludes zero-padded contacts. Attention and feed-forward sub-blocks each include a residual connection and post-normalization.

The Transformer comprised three stacked attention blocks with an embedding dimension of 32 and 8 attention heads. Within each block, the contact embeddings were linearly projected into query (*Q*), key (*K*), and value (*V*) representations. For attention head ℎ, the unnormalized attention score between contacts *i* and *j* was calculated as

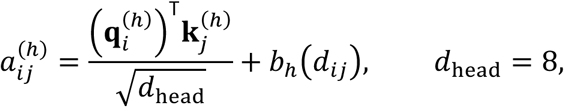

where *d_i_*_j_ is the Euclidean distance between the contacts’ MNI coordinates, and *b*_ℎ_(*d_i_*_j_) is a learned, head-specific distance bias. The distance bias was generated independently within each attention block by a two-layer neural network,

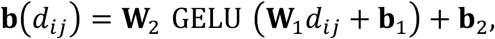

with dimensions 1 → 16 → 4, producing one bias value for each attention head. The distance bias was added to the scaled dot-product attention logits before masking and softmax normalization. This allowed the contribution of one contact to another to vary as a learned, non-parametric function of their physical separation.

Each attention layer was followed by a residual connection and layer normalization. The resulting embeddings were passed through a feed-forward network with dimensions 32 → 64 → 32, Gaussian error linear unit (GELU) activation after both linear layers, and dropout of 0.1. A second residual connection and layer-normalization operation completed each block. The architecture, therefore, used post-normalization, with layer normalization applied after each residual addition.

A shared linear output layer mapped the final 32-dimensional embedding of each contact to two logits representing the non-target and target classes. Softmax normalization was applied outside the model to obtain the predicted probability that each contact belonged to the resected tissue.

### Model training, validation and ensemble construction

Three models were generated using leave-one-public-cohort-out development. In each fold, patients from two of the three public cohorts were used for model fitting, while the third cohort was held out in its entirety for validation and checkpoint selection. Cohorts were rotated manually across the three folds so that each served once as the validation cohort. No additional patient-level split was performed within either the fitting or validation cohorts. Because the held-out public cohort was used to select the checkpoint, it constituted a validation set rather than an independent test set. Splitting was performed at the patient level: all segments and contacts from a given patient were assigned to the same fold.

During each training epoch, one recording segment (10 minute) was sampled randomly from each patient in the fitting cohorts. During validation, predictions were generated for every available segment. Segment-level probabilities were subsequently grouped by patient and contact and averaged across segments to obtain one probability per contact. The three checkpoints selected across the development folds were fixed before external evaluation. At external inference, each checkpoint was applied independently, and the three resulting contact-level probabilities were combined using an unweighted arithmetic mean. No external-cohort data were used for model fitting, hyperparameter selection, checkpoint selection, ensemble weighting or probability recalibration.

Network parameters were optimized for 50 epochs using the Adam optimizer^48^ with a fixed learning rate of 0.0025 and a mini-batch size of four patients, i.e. (B=4, CH=256, F=36). Default Adam parameters were used (β₁ = 0.9, β₂ = 0.999 and ε = 10⁻⁸), without weight decay or a learning-rate scheduler. No early stopping, gradient clipping, or mixed-precision training was applied. To improve tolerance to missing or noisy inputs applied Dropout with p=0.25 during training.

The principal classification loss was weighted two-class cross-entropy with label smoothing of 0.149. The positive class was weighted threefold relative to the negative class to address contact-level class imbalance. In addition, losses from patients with favorable surgical outcomes were weighted threefold relative to those from other patients. Only non-padded contacts contributed to the loss. Two regularization terms were added to the classification loss. A spread loss, with a coefficient of 1.0, penalized the difference between the maximum and minimum predicted probabilities within a patient when this range was smaller than one. Minimizing this term encouraged greater separation between contact-level predictions and reduced output collapsing toward uniform probabilities across an implantation. After epoch 30, a self-distillation term was introduced. This term calculated the Kullback–Leibler divergence between the current model’s softmax output distribution and the fixed output distribution of the best validation checkpoint obtained up to that point. It therefore discouraged the model from deviating substantially from its best previously observed validation state, promoting local neighbourhood search.

After every epoch, predictions were generated for all segments in the held-out validation cohort and averaged by patient and contact. Patient-level AUPRC and AUROC were then calculated. The best model state was retained in memory during training and saved after completion of all 50 epochs. One model was trained for each of the three public cohort rotations, producing three training runs in total, forming classification ensamble.

### Model evaluation

The primary performance measures were AUPRC and AUROC, which have been recommended as complementary metrics for evaluating contact-level localization of the EZ^50^. Both metrics were calculated separately for each patient by comparing contact-level ensemble probabilities with binary resection labels using the *average_precision_score* and *roc_auc_score* functions from scikit-learn. Thus, each patient contributed one AUPRC and one AUROC value, and all patients were weighted equally in cohort-level summaries. Metrics were summarized across patients using the median and interquartile range (IQR) and stratified by postoperative outcome and institution. AUPRC was emphasized because the proportion of resected contacts varied among patients. The chance-level AUPRC was calculated separately for each patient as the proportion of analyzed contacts that were resected, whereas the chance-level AUROC was 0.5.

Interictal spike rate, the clinically annotated SOZ, and the SOZ expanded by a 10-mm Euclidean margin (SOZ+10 mm) were evaluated as contact-level reference measures. For each patient, spike rates were summed across all available feature windows for each contact. SOZ was represented as a binary contact-level indicator. For SOZ+10 mm, all contacts located within a Euclidean distance of 10 mm or less from any SOZ contact were assigned a positive value. AUROC and AUPRC were calculated using the same resection labels as for the model. If a reference measure had only one unique value within a patient, its AUPRC and AUROC were treated as undefined.

For operating-point analyses, model scores were normalized relative to the highest predicted probability within each patient. Contacts with an ensemble probability greater than *t* × *p_max_*, where *t* was 0.75, 0.80, 0.85, or 0.90, and *p_max_*was the maximum probability for that patient, were designated as suprathreshold seed contacts. The selected region was then expanded to include every implanted contact within 3, 6, 9, or 12 mm of any seed contact. Sensitivity, specificity, and positive predictive value were calculated separately for each patient from the resulting binary contact classifications.

Spatial prediction–resection displacement was quantified as the minimum Euclidean distance in MNI space between the contact receiving the highest ensemble probability and any resected contact. A distance of 0 mm indicated that the highest-scoring contact was itself resected. If multiple contacts shared the maximum probability, the first contact in the patient-level prediction table was selected.

### Statistical analysis

The patient was the independent statistical unit. Continuous variables and performance measures were summarized as medians and IQRs. AUPRC, AUROC, and peak-prediction distance were compared between independent Engel I and Engel II–IV groups using two-sided Mann–Whitney U tests; rank-biserial correlation was reported as an effect size, with positive values indicating higher performance or the prespecified direction in Engel I. Rank-biserial correlation (*r_rb_*) was reported as the effect size.

Comparisons of the model with spike rate, SOZ, and SOZ+10 mm were restricted to Engel I patients and performed using two-sided paired Wilcoxon signed-rank tests. Each comparison included only patients with non-missing values for both methods. To account for the three baseline comparisons, *P* values were adjusted using the Benjamini–Hochberg false-discovery-rate procedure separately for AUPRC and AUROC. Adjusted *P* values are reported as *P_FDR_*.

Institution-specific analyses were treated as subgroup analyses and reported alongside the pooled external analysis. Motol was excluded from between-outcome tests because it contained no Engel II–IV patients. Statistical significance was assessed at α = 0.05. Analyses were performed in R version 4.5.3 using *dplyr* version 1.2.1 and in Python version 3.11.4 using *scikit-learn* version 1.8.0.

### Selection of published studies for comparison

We selected published studies of automated multifeatured localization of epileptogenic tissue from iEEG for contextual comparison. Studies using datasets partially overlapping with ours were included if they combined multiple features for localization. Studies using independent datasets were identified among those included in a published systematic review by Hrtonova et al^33^. These studies were eligible if they (i) combined multiple features for localization, (ii) evaluated performance using cross-validation with separation of training and test data, (iii) included at least 15 patients in performance evaluation, and (iv) reported AUROC or AUPRC. Evaluation targets and surgical outcome groups were recorded alongside performance metrics to clarify differences between studies. These comparisons provide context rather than a direct benchmark because evaluation targets, cohorts, and validation procedures differ.

### Missing data, study size and bias safeguards

Analyses used available cases; no imputation was performed. Patients lacking a variable required for a specific comparison were excluded only from that analysis, and the corresponding denominator was reported. For example, SOZ annotations were unavailable for three Engel I patients in the Brno cohort, resulting in n = 117 for pooled SOZ-based comparisons. The study size was determined by all patients meeting eligibility and data-availability criteria in the selected public resources and participating external cohorts; no formal prospective sample-size calculation was performed.

Several design measures were used to limit bias. Public development and private evaluation cohorts were completely separated, and external predictions were generated without retraining or recalibration. Cohort eligibility, signal selection, electrode and resection labeling, and outcome ascertainment were completed before model application and without knowledge of its predictions. Performance was calculated within patients to avoid treating contacts as independent observations, and results were stratified by postoperative outcome because resected tissue is an imperfect proxy for epileptogenic tissue, particularly in patients with persistent seizures. Preprocessing and measurement procedures were not retrospectively harmonized across cohorts, reflecting differences in routine clinical practice and data acquisition. The model was intentionally evaluated without adjustment for center, implantation type, age, or recording state to assess its performance under this real-world heterogeneity.

## Data and code availability

The public development datasets are available through OpenNeuro under the accession numbers listed above. The code will be made publicly available upon journal acceptance. The external validation datasets are available from the principal investigators of the contributing institutions upon reasonable request, subject to institutional data-sharing agreements.

## Acknowledgements

Ministry of Health of the Czech Republic, project NW25-08-00212; Canadian Institutes of Health Research funding (PJT-175056), Infrastructure Funding of the American Epilepsy Society, Start-up funding of Duke University.

Pilar Bosque-Varela received travel grants and honoraria from UCB as well as a grant from the Austrian Epilepsy Society (ÖGfe) for her 6-month research fellowship at Duke Medical Hospital.

Radek Janča, Petr Ježdík were financed by EU-Next Generation EU; ERDF-Project Brain Dynamics (No.CZ.02.01.01/00/22 008/0004643)

## Supplementary figures

**Supplementary Figure 1.**
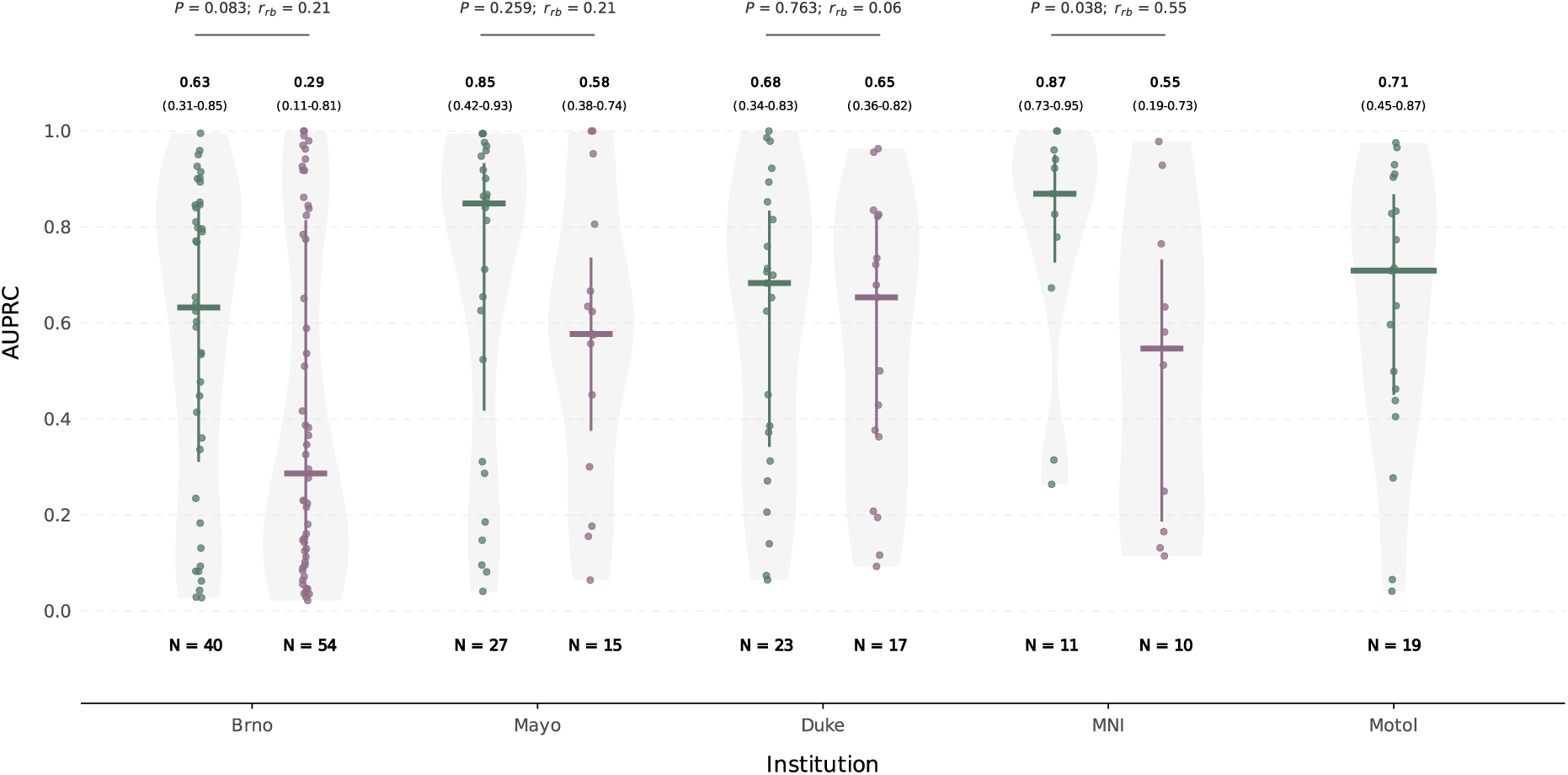
Institution-specific AUPRC stratified by postsurgical outcome. Distribution of patient-level AUPRC values for the iEEG Transformer ensemble in each external validation cohort, stratified by seizure-free (Engel I; green) and non-seizure-free outcomes (Engel II–IV; purple). Points represent individual patients, horizontal bars indicate medians, vertical bars indicate interquartile ranges, and shaded violin plots show the distributions. Values above each group report the median (interquartile range); sample sizes are shown below. Differences between outcome groups were assessed within each institution using a two-sided Mann–Whitney U test, with the prespecified alternative hypothesis that AUPRC was higher in Engel I than in Engel II–IV. Effect sizes are reported as rank-biserial correlations (*r*_rb_), with positive values indicating higher AUPRC in Engel I. The Motol cohort included only Engel I patients and was therefore not included in the outcome-group comparison. AUPRC, area under the precision–recall curve.

**Supplementary Figure 2.**
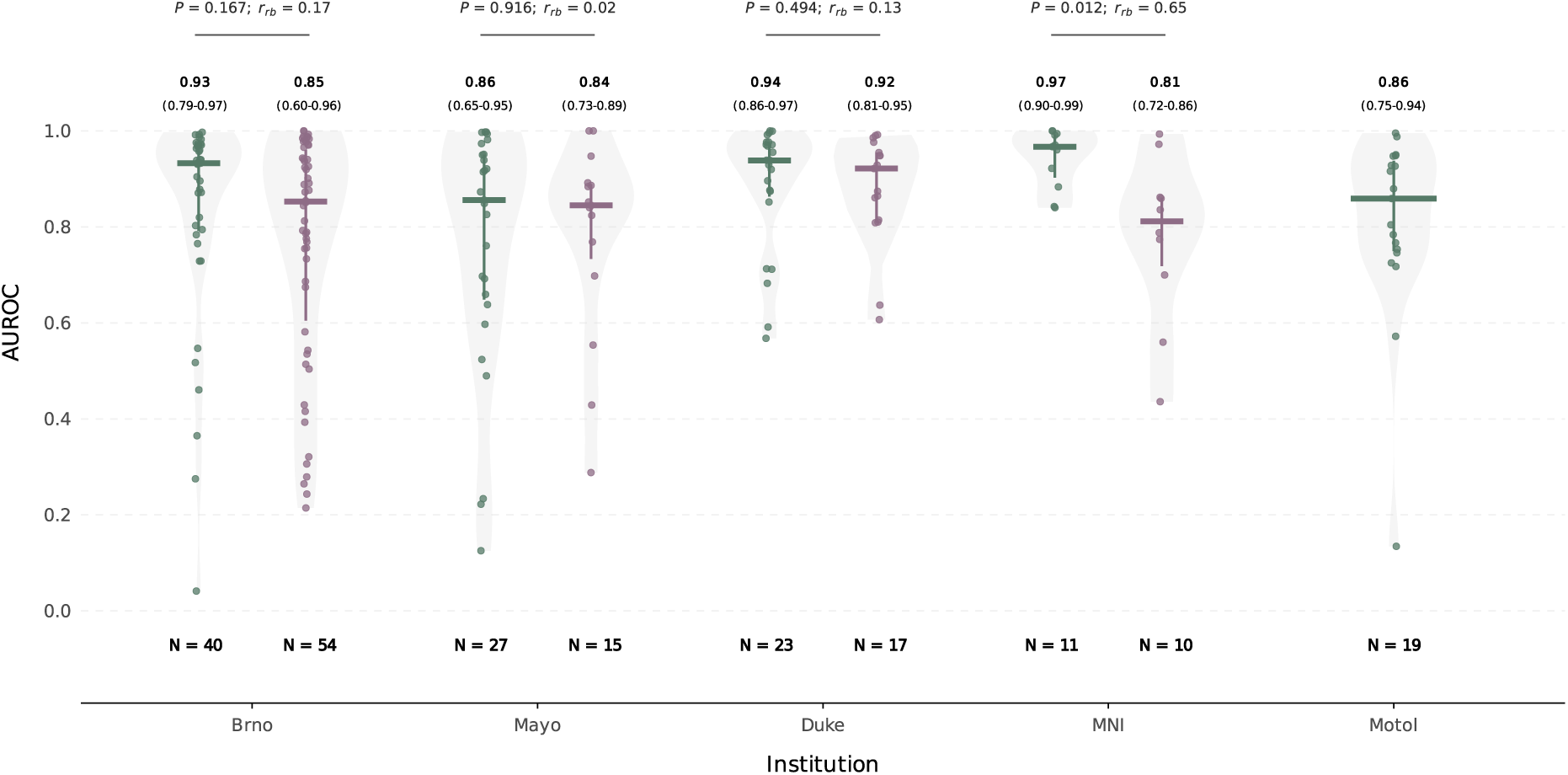
Institution-specific AUROC stratified by postsurgical outcome. Distribution of patient-level AUROC values for the iEEG Transformer ensemble in each external validation cohort, stratified by seizure-free (Engel I; green) and non-seizure-free outcomes (Engel II–IV; purple). Points represent individual patients, horizontal bars indicate medians, vertical bars indicate interquartile ranges, and shaded violin plots show the distributions. Values above each group report the median (interquartile range); sample sizes are shown below. Differences between outcome groups were assessed within each institution using a two-sided Mann–Whitney U test, with the prespecified alternative hypothesis that AUROC was higher in Engel I than in Engel II–IV. Effect sizes are reported as rank-biserial correlations (*r*_rb_), with positive values indicating higher AUROC in Engel I. The Motol cohort included only Engel I patients and was therefore not included in the outcome-group comparison. AUROC, area under the receiver operating characteristic curve.

**Supplementary Figure 3.**
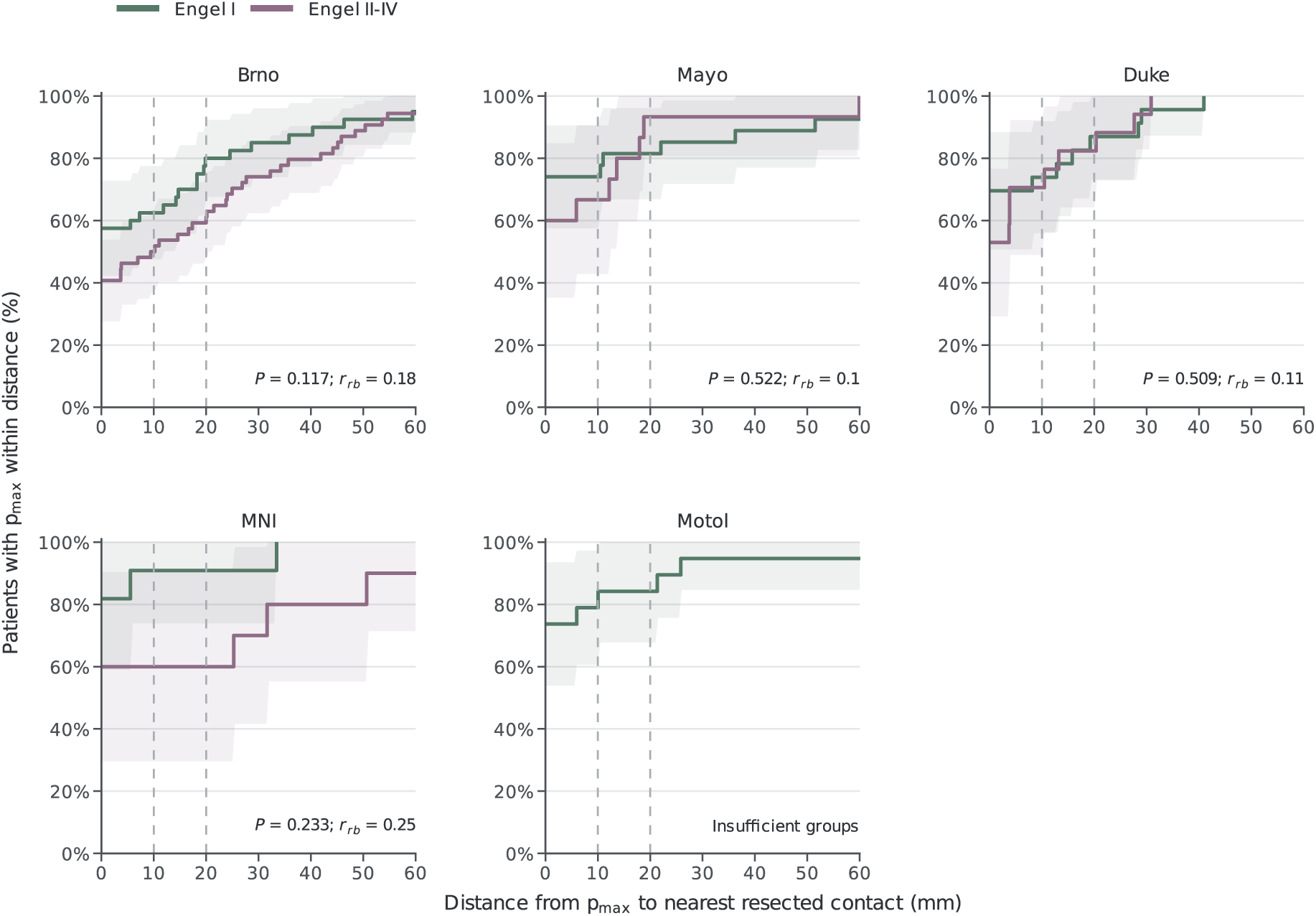
Institution-specific spatial displacement of peak model predictions relative to resected tissue. Empirical cumulative distributions of the Euclidean distance (*d*) between the contact assigned the highest model probability (*p*_max_) and the nearest resected contact, shown separately for each external cohort and stratified by postsurgical outcome. At each distance, the y-axis indicates the proportion of patients whose *p*_max_ contact was located within that distance of a resected contact; *d =* 0 mm indicates that the contact was resected. Shaded bands indicate 95% confidence intervals, and vertical dashed lines mark distances of 10 and 20 mm. Differences between Engel I and Engel II–IV patients were assessed using two-sided Mann–Whitney-tests. No institution-specific comparison reached statistical significance. Motol included only patients with Engel I outcomes and was therefore not tested. Sample sizes are indicated in each panel.

## Supplementary tables

**Supplementary Table 1.**
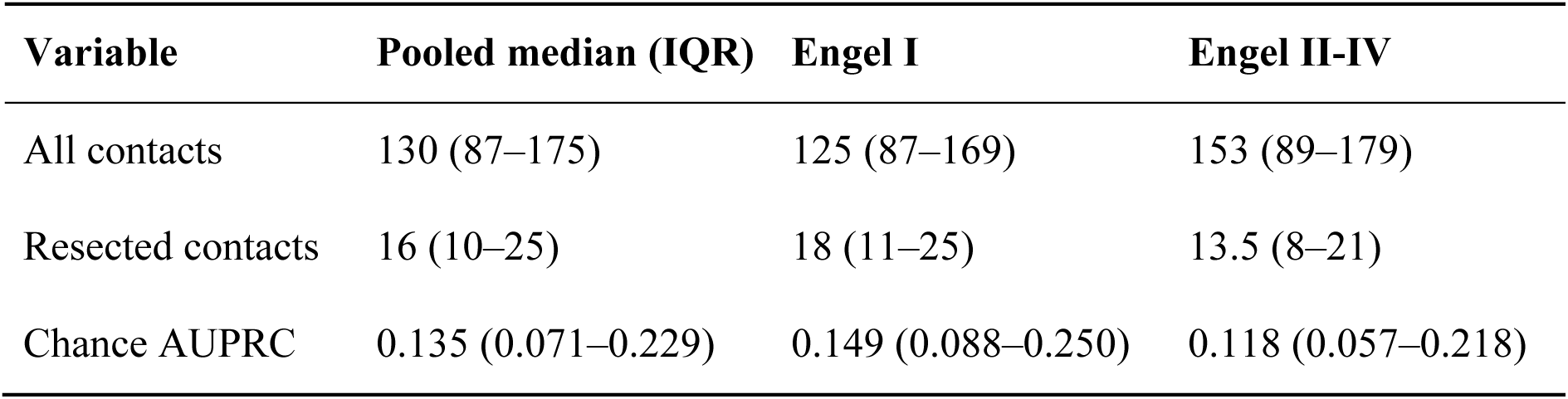
Contact counts and chance AUPRC reference levels by postsurgical outcome. Values are medians (interquartile ranges) across patients, reported for the pooled cohort (n = 216), Engel I patients (n = 120), and Engel II–IV patients (n = 96). Chance AUPRC was calculated separately for each patient as the number of resected contacts divided by the total number of evaluated contacts. AUPRC, area under the precision–recall curve.

**Supplementary Table 2.** Pooled and institution-specific comparison of the Transformer ensemble with interictal and clinical localization baselines. Data are medians with interquartile ranges (IQRs) shown on the second line, for the pooled external evaluation sample and by cohort and postsurgical outcome. Model, Transformer ensemble; Spike rate, interictal spike rate; SOZ, clinically annotated seizure-onset zone; SOZ+10 mm, SOZ expanded by a 10-mm Euclidean margin. *For SOZ-based analyses, pooled n = 117 for Engel I and n = 79 for Engel II–IV; in Brno, n = 37 in each outcome group because SOZ annotations were unavailable for a subset of patients. Motol included only patients with Engel I outcomes. AUROC, area under the receiver operating characteristic curve; AUPRC, area under the precision–recall curve.

| Cohort | Outcome | n | Model |  | Spike rate |  | SOZ |  | SOZ+10 mm |  |
|---|---|---|---|---|---|---|---|---|---|---|
|  |  |  | AUROC | AUPRC | AUROC | AUPRC | AUROC | AUPRC | AUROC | AUPRC |
| Pooled | I | 120 | 0.92 (0.76–0.97) | 0.71 (0.37–0.90) | 0.80 (0.68–0.92) | 0.55 (0.34–0.75) | 0.63 (0.56–0.75)* | 0.41 (0.26–0.55)* | 0.72 (0.61–0.87)* | 0.45 (0.30–0.61)* |
|  | II-IV | 96 | 0.86 (0.70–0.95) | 0.42 (0.15–0.81) | 0.75 (0.63–0.88) | 0.33 (0.15–0.59) | 0.65 (0.55–0.81)* | 0.36 (0.22–0.51)* | 0.74 (0.58–0.92)* | 0.36 (0.26–0.54)* |
| Brno | I | 40 | 0.93 (0.79–0.97) | 0.63 (0.31–0.85) | 0.83 (0.72–0.93) | 0.48 (0.34–0.76) | 0.64 (0.58–0.89)* | 0.42 (0.28–0.79)* | 0.71 (0.63–0.94)* | 0.45 (0.34–0.60)* |
|  | II-IV | 54 | 0.85 (0.60–0.96) | 0.29 (0.11–0.81) | 0.79 (0.67–0.88) | 0.32 (0.12–0.64) | 0.73 (0.61–0.90)* | 0.41 (0.26–0.61)* | 0.78 (0.68–0.97)* | 0.37 (0.29–0.70)* |
| Mayo | I | 27 | 0.86 (0.65–0.95) | 0.85 (0.42–0.93) | 0.69 (0.54–0.82) | 0.62 (0.32–0.76) | 0.56 (0.52–0.68) | 0.46 (0.31–0.54) | 0.63 (0.52–0.71) | 0.48 (0.29–0.57) |
|  | II-IV | 15 | 0.84 (0.73–0.89) | 0.58 (0.38–0.74) | 0.64 (0.52–0.73) | 0.45 (0.19–0.67) | 0.57 (0.53–0.67) | 0.34 (0.21–0.43) | 0.58 (0.53–0.74) | 0.34 (0.21–0.44) |
| Duke | I | 23 | 0.94 (0.86–0.97) | 0.68 (0.34–0.83) | 0.86 (0.75–0.92) | 0.53 (0.35–0.66) | 0.63 (0.57–0.75) | 0.26 (0.17–0.47) | 0.75 (0.60–0.88) | 0.30 (0.21–0.51) |
|  | II-IV | 17 | 0.92 (0.81–0.95) | 0.65 (0.36–0.82) | 0.76 (0.66–0.88) | 0.36 (0.21–0.59) | 0.57 (0.53–0.77) | 0.27 (0.20–0.43) | 0.63 (0.56–0.87) | 0.31 (0.21–0.54) |
| MNI | I | 11 | 0.97 (0.90–0.99) | 0.87 (0.73–0.95) | 0.91 (0.61–0.97) | 0.64 (0.23–0.80) | 0.74 (0.68–0.78) | 0.42 (0.32–0.56) | 0.83 (0.76–0.91) | 0.49 (0.34–0.63) |
|  | II-IV | 10 | 0.81 (0.72–0.86) | 0.55 (0.19–0.73) | 0.68 (0.52–0.73) | 0.27 (0.22–0.38) | 0.69 (0.64–0.81) | 0.40 (0.30–0.44) | 0.78 (0.70–0.89) | 0.48 (0.30–0.51) |
| Motol | I | 19 | 0.86 (0.75–0.94) | 0.71 (0.45–0.87) | 0.80 (0.72–0.86) | 0.63 (0.39–0.74) | 0.63 (0.60–0.69) | 0.40 (0.31–0.53) | 0.75 (0.71–0.82) | 0.49 (0.35–0.65) |

**Supplementary Table 3.**
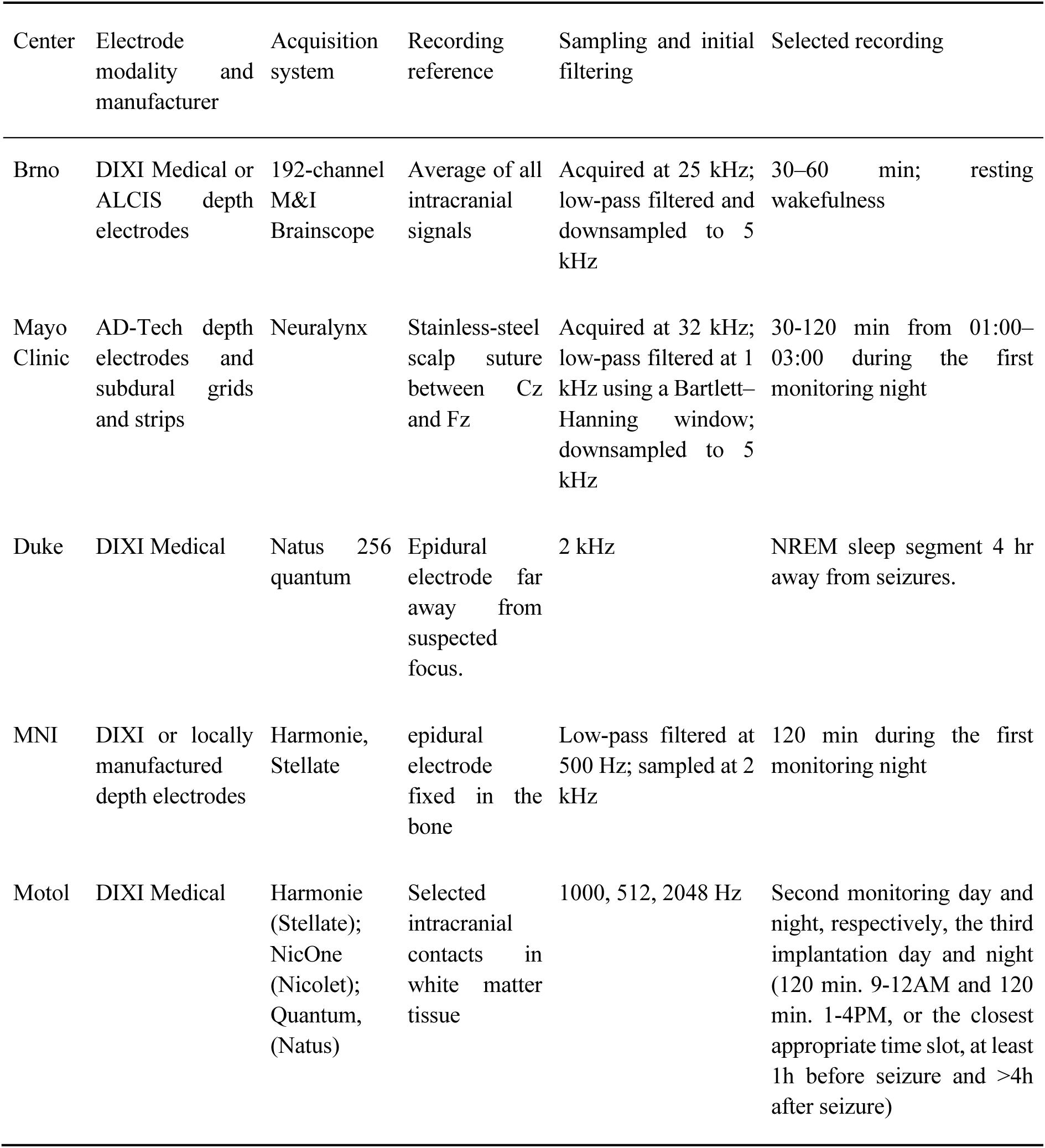
Center-specific iEEG acquisition and data-selection procedures. Acquisition settings refer to the recordings included in this study. Where procedures differed across patients within a center, the applicable range or configurations are reported. Abbreviations: iEEG, intracranial electroencephalography; MNI, Montreal Neurological Institute; sEEG, stereo-electroencephalography.

